# Understanding the interrelationships of type-2 diabetes and hypertension with brain and cognitive health: a UK Biobank study

**DOI:** 10.1101/2021.11.17.21266262

**Authors:** Danielle Newby, Victoria Garfield

## Abstract

**Aims/hypothesis:** Diabetes and hypertension are associated with poorer cognitive and brain health. Less is known about comorbid diabetes and hypertension and: cognitive and brain health in mid-life. We hypothesised that individuals with both diabetes and hypertension have worse cognitive and brain health.

**Methods:** We used the UK Biobank cohort, a population-based study which recruited 500,000 individuals, aged 40-69 years. Type-2 diabetes was assigned using self-report, HbA1c and clinical data, while hypertension was defined based on self-report. Our outcomes included a breadth of brain structural magnetic resonance imaging (MRI) parameters and cognitive function tests in a maximum of 38918 individuals. We tested associations in a cross sectional design firstly between those with diabetes (n = 2043) versus those without (reference category, n = 36875) then we tested associations between comorbid diabetes/hypertension (reference category, n=1283) and our outcomes by comparing this group with those with only diabetes (n=760), hypertension (n=9649) and neither disease (n=27226). Our analytical approach comprised linear regression models, with adjustment for a range of demographic and health factors.

**Results:** Those with diabetes had worse overall brain health, as indexed by multiple neuroimaging parameters, with the exception of gFA (white matter integrity) and the amygdala. The largest difference was observed in the pallidum (β=0.179, 95%CI=0.137;0.220). Individuals with diabetes had poorer performance on certain cognitive tests, with the largest difference observed in the symbol digit substitution test (β=0.132, 95%CI=0.079;0.187). Compared to individuals with comorbid diabetes and hypertension, those with only hypertension had better brain health overall, with the largest difference observed in the pallidum (β=0.189, 95%CI=0.241;0.137), while those with only diabetes differed in total grey volume (β=0.150, 95%CI=0.122;0.179). Compared with individuals who had comorbid diabetes and hypertension, those with only diabetes performed distinctly better on the verbal and numeric reasoning task (β=0.129, 95%CI=0.077;0.261), whereas those with only hypertension performed better on the symbol digit substitution task (β=0.117, 95%CI=0.048;0.186)

**Conclusions/interpretation:** Individuals with comorbid diabetes and hypertension have worse brain and cognitive health compared to those with only one of these diseases. These findings potentially suggest that prevention of both diabetes and hypertension may delay changes in brain structure, as well as cognitive decline and dementia diagnosis.

**Research in context:** *What is already known about this subject?:*

- Diabetes and hypertension are both independently associated with poorer brain health, cognition and increase in dementia risk
- 50% of adults with diabetes also have hypertension
- Comorbid diabetes and hypertension are associated with excess dementia risk

*What is the key question?:*

- Do individuals with both diabetes and hypertension have worse cognitive and brain health?

*What are the new findings?:*

- Individuals with diabetes have poorer brain health and cognitive performance using a breadth of neuroimaging measures and cognitive tests
- Those with both diabetes and hypertension have worse brain health and cognitive performance (particularly processing speed) compared to those with only hypertension

*How might this impact on clinical practice in the foreseeable future?:*

- Prevention of both diabetes and hypertension may help delay changes in brain structure and cognitive decline as well as reduce future dementia risk.

## INTRODUCTION

Type-2 diabetes is associated with excess risk of dementia [1, 2], worse cognitive function [3, 4] and changes in brain structure and function [1]. Large-scale population based studies, such as the UK Biobank (UKB), have recently expanded our understanding of the relationship between diabetes, cognitive function and brain health (as captured by magnetic resonance imaging (MRI) [5–7]). Important insights from these studies include that diabetes is associated with: slower reaction times [5, 7], poorer brain health as indexed by structural and diffusion indices and increased risk of vascular, Alzheimer’s and all-cause dementia [6, 7]. The relationship between hypertension and a range of adverse cognitive and brain health parameters is also well established [8–12]. Hypertension in mid-life is also known to increase the risk of both Alzheimer’s and vascular dementia [10, 12].

Importantly, global estimates show that more than 50% of adults with diabetes also have a hypertension diagnosis [13]. Epidemiological data suggest that those with both diabetes and hypertension have a 23% excess risk of dementia, in comparison with individuals with diabetes but no hypertension whose excess risk is around 19%. Also, evidence from the UKB suggests that the additive combination of diabetes and hypertension is related to worse cognitive performance when accounting for a number of confounding factors [5].

As described above, there have been recent important advances in how diabetes and hypertension are associated with cognitive and brain health, yet a number of evidence gaps remain to be filled. To date, no research has investigated: whether comorbid diabetes and hypertension is associated with worse brain health, as captured by multiple structural neuroimaging outcomes; nor have they used a breadth of measures to define diabetes in this context. In this study, we used data from the UKB to test two a priori hypotheses: i) participants with diabetes have poorer brain health and cognitive function compared to those without diabetes and ii) participants with diagnosed diabetes and hypertension have poorer brain health and cognitive function than those with only one of these diseases or with neither of these diseases. We tested both of our hypotheses using multiple neuroimaging outcomes and cognitive tests to capture brain health and cognitive function.

## METHODS

### Study design

A cross sectional study of 38918 participants with brain MRI data in UK Biobank was used to determine the association between diabetes and those with either hypertension and/or diabetes with several global brain volumes, grey subcortical, white matter micro and macrostructures volumes and cognitive tests.

### Setting

UK Biobank is a large prospective cohort of over half a million participants. All participants, aged between 40 and 69, initially attended baseline assessment visits from 2006 to 2010 where they completed a series of physical, sociodemographic, cognitive and medical assessments [14]. Follow up visits have taken place including typical assessments as with baseline visit but with whole body imaging including MRI brain imaging. To date nearly 50K participants have MRI brain imaging carried with over 40K participants with data currently available. These participants also completed the same battery of assessments as the baseline visit. UK Biobank received ethical approval from the Research Ethics Committee (11/NW/0382). Volunteers gave informed consent for their participation.

### Participants

Participants who attended the assessment centre for an MRI brain scan were included in this study. These participants also provided demographic, health, and socioeconomic information using touchscreen questionnaires as well as taking part in a nurse-led interview asking questions about medical history and medications. Participants who reported they had any neurodegenerative or related diseases were excluded from this analysis (n = 612) as in previous work [9]. A full list of these diseases and UK Biobank field codes for all variables used in this work can be found in **Supporting Tables 1 and 2**. We additionally removed participants with any diagnosis of either type I (n = 152), gestational diabetes (n = 4) or unspecified diabetes (n = 6) after defining our diabetes population. This resulted in 38918 participants included in our analysis

### Variables

#### Exposures: Diabetes and hypertension

Participants were defined with diabetes using information on self-reported diagnosis, self-reported medications, biochemistry and clinical data. Participants were defined with hypertension based on self-reported diagnosis and self-reported medications. The UK Biobank field codes to define diabetes and hypertension used in this work and further information regarding definitions of phenotypes can be found in the **Supporting Information**.

#### Outcome: neuroimaging

Brain MRIs were acquired on a Siemens Skyra 3 T scanner with a standard Siemens 32-channel head coil. We utilised Imaging Derived Phenotypes (IDPs) derived from the raw brain MRI images which were generated using an image-processing pipeline developed and quality controlled centrally by UK Biobank [15]. In this work, we included total brain volume, grey matter volume, white matter hyperintensity (WMH) volume and ventricular CSF. We also analysed subcortical volumes (accumbens, amygdala, caudate, hippocampus, pallidum, putamen, and thalamus) by averaging left and right measures and latent measures of tract-averaged fractional anisotropy (FA) and mean diffusivity (MD) of all the white matter tracts. FA and MD are two metrics imaged with diffusion-tensor imaging (DTI) indicative of white matter tract microstructural integrity: higher FA values suggest better health, whereas higher MD suggests worse white matter tract health. Due to the high correlation of the white matter microstructural properties across the brain of individual regions of FA and MD, single general latent measures of FA (*g*FA) and MD (*g*MD) were created using confirmatory factor analysis as described previously [6, 16]. Due to the low correlation of the subcortical volumes (**Supporting Information Figure S1**) we did not create latent measures for these brain measures. Outlier data points, defined as being further than ±4 SD from the mean, were excluded (<1% of values).

#### Outcome: cognitive function

Cognitive function was assessed using the cognitive tests verbal–numerical reasoning, pairs matching (memory), reaction time (processing speed), Matrix Pattern (nonverbal reasoning), Symbol-Digit Substitution (processing speed/executive function), tower rearranging (executive function/ planning), and the difference between Trail-Making Tests (TMT) B and A (processing speed/executive function). Further information regarding the cognitive tests can be found https://biobank.ctsu.ox.ac.uk/crystal/label.cgi?id=100026. Verbal–numerical reasoning, pairs matching and reaction time cognitive tests were bespoke to UK Biobank. The remaining cognitive tests are validated tests, which were additionally administered at the imaging visits from 2016. Higher values indicate better cognitive performance on verbal and numeric reasoning, matrix reasoning, symbol-digit substitution, and tower rearranging, and worse cognitive performance on the reaction time, pairs matching, and TMT B - TMT A.

#### Covariates

In this study we selected the following covariates for adjustment in our models: demographic measures (age + sex + deprivation + educational attainment + ethnicity), as well as head size and MRI scanner position variables (for neuroimaging outcomes only) and standard cardiovascular risk factor measures (smoking + BMI + hypertension + high cholesterol).

Age at assessment was measured in whole years and sex was self-reported as male or female. Educational qualifications were self-reported and were dichotomized into whether participants held a university/college degree or not. Self-reported ethnicity was dichotomized into white or non-white and if was missing was obtained from the baseline assessment visit.

Assessment centre was a multilevel variable of the assessment centres utilised for the repeated imaging visits (n = 3). Townsend deprivation index was calculated before the baseline visit and was split into quintiles. BMI was calculated from valid height (cm) and weight (kg) measurements obtained by UK Biobank and was used as a continuous measure. Smoking status was self-reported and categorized into never smoked, current or former smoker. For hyperlipidemia a combination of self-reported and medication records were used. Where participants responded ‘Do not know’ or ‘Prefer not to answer’ these were treated as missing (<1%) and missing data was not imputed.

### Statistical methods

All analyses were performed using R version 4.0.2. Descriptive statistics were generated to characterise the study cohort at baseline.

#### Modelling approach

Natural Log transformations were applied to reaction time and WMH. For the pairs matching cognitive test a natural log transformation + 1 (ln(x+1)) was used and for TMT B – A, a square root transformation was applied to make these outcomes normally distributed prior to analysis. All other outcome variables were normally distributed. Standardised beta coefficients are reported for all analyses to facilitate comparison of associations across the neuroimaging and cognitive outcomes. We converted all of our betas so that they would be in the same direction in order to aid interpretability. Therefore, all standardised betas with a positive value indicate better brain health or cognitive performance and negative values indicate poorer brain health and cognitive performance. For the neuroimaging outcomes, WMH, Vascular CSF, gMD results were converted to the same direction as all other neuroimaging measures and for the cognitive outcomes, reaction time, pairs matching and Trail making test B-A results were converted to the same direction as all the other cognitive tests.

In stage 1 of our analyses we tested our first a priori hypothesis and compared brain and cognitive function measures between those with (n = 2043) and without diabetes (n = 36875). In stage 2, we tested our second a priori hypothesis, that comorbid diabetes and hypertension would associate more strongly with worse brain and cognitive health. Thus, we compared outcomes across those with both hypertension and diabetes (n = 1283) versus those with only diabetes (n = 760), only hypertension (n = 9649) or no hypertension/diabetes (n = 27226). We fitted two linear regression models for stages 1 and 2 of our analyses; the first was adjusted for demographics (model 1) and the second was additionally adjusted for cardiovascular related covariates, as described earlier (model 2).

## RESULTS

### Sample characteristics

A total of 38918 individuals were included in the study, of whom 36875 did not have diabetes and 2043 had known diabetes. Those with diabetes were more likely to be male, not have a degree, not of caucasian ethnicity, be current smokers, reside in the most deprived quintile, have a higher BMI and had the highest prevalence of hypercholesterolaemia and hypertension (**Table 1**).

**Table 1:**
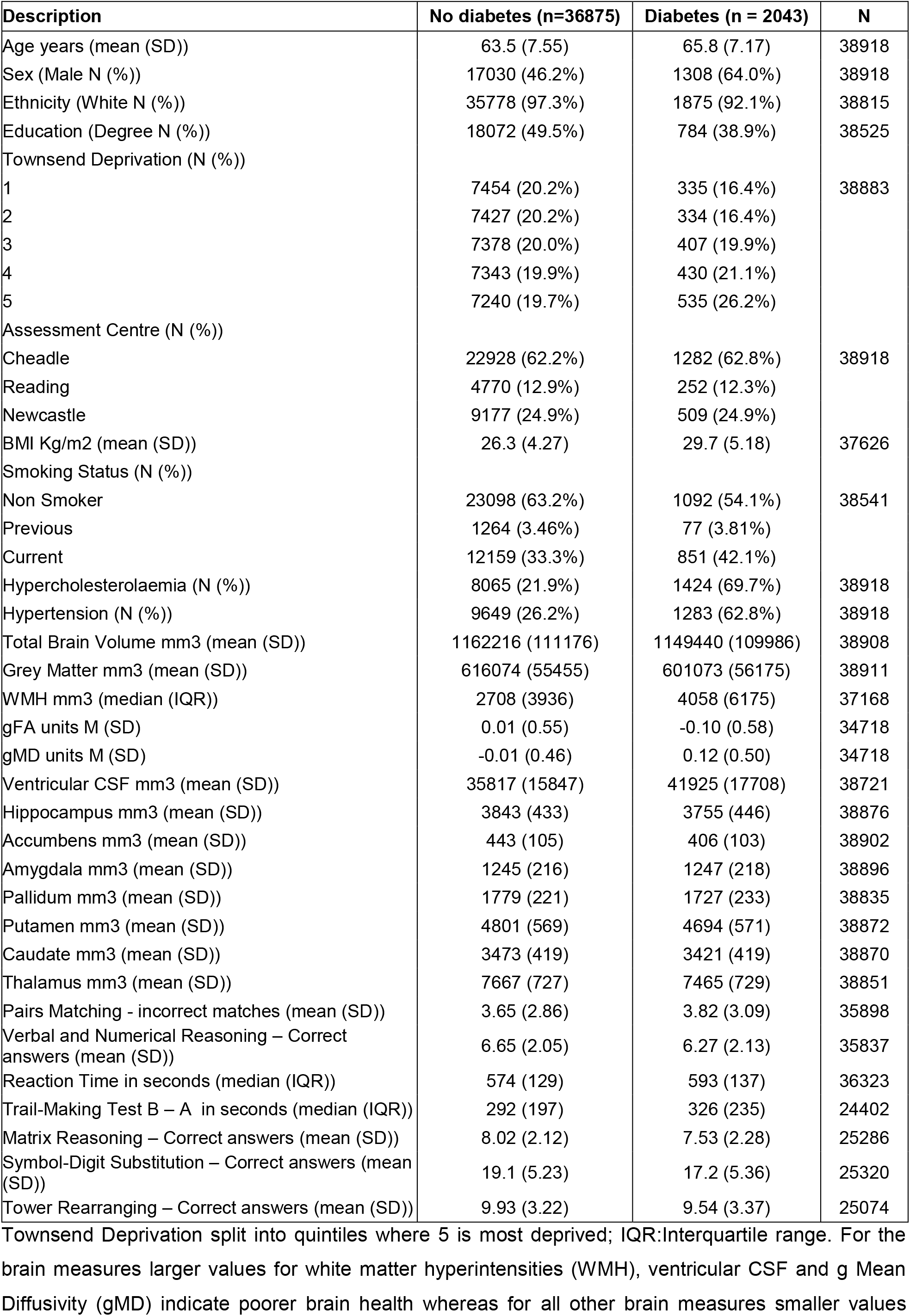
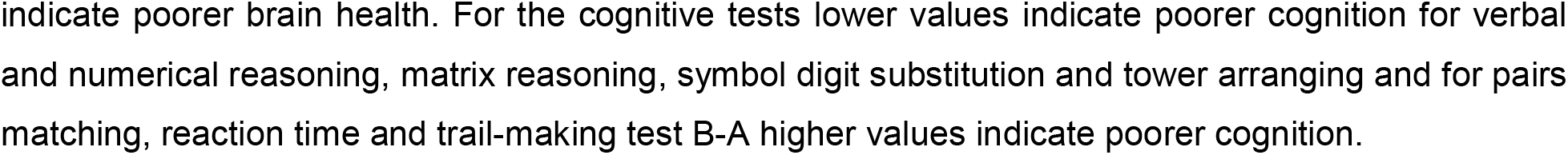
Characteristics of UK Biobank participants at imaging visit stratified by diabetes diagnosis.

| Description | No diabetes (n=36875) | Diabetes (n = 2043) | N |
| --- | --- | --- | --- |
| Age years (mean (SD)) | 63.5 (7.55) | 65.8 (7.17) | 38918 |
| Sex (Male N (%)) | 17030 (46.2%) | 1308 (64.0%) | 38918 |
| Ethnicity (White N (%)) | 35778 (97.3%) | 1875 (92.1%) | 38815 |
| Education (Degree N (%)) | 18072 (49.5%) | 784 (38.9%) | 38525 |
| Townsend Deprivation (N (%)) |  |  |  |
| 1 | 7454 (20.2%) | 335 (16.4%) | 38883 |
| 2 | 7427 (20.2%) | 334 (16.4%) |  |
| 3 | 7378 (20.0%) | 407 (19.9%) |  |
| 4 | 7343 (19.9%) | 430 (21.1%) |  |
| 5 | 7240 (19.7%) | 535 (26.2%) |  |
| Assessment Centre (N (%)) |  |  |  |
| Cheadle | 22928 (62.2%) | 1282 (62.8%) | 38918 |
| Reading | 4770 (12.9%) | 252 (12.3%) |  |
| Newcastle | 9177 (24.9%) | 509 (24.9%) |  |
| BMI Kg/m2 (mean (SD)) | 26.3 (4.27) | 29.7 (5.18) | 37626 |
| Smoking Status (N (%)) |  |  |  |
| Non Smoker | 23098 (63.2%) | 1092 (54.1%) | 38541 |
| Previous | 1264 (3.46%) | 77 (3.81%) |  |
| Current | 12159 (33.3%) | 851 (42.1%) |  |
| Hypercholesterolaemia (N (%)) | 8065 (21.9%) | 1424 (69.7%) | 38918 |
| Hypertension (N (%)) | 9649 (26.2%) | 1283 (62.8%) | 38918 |
| Total Brain Volume mm3 (mean (SD)) | 1162216 (111176) | 1149440 (109986) | 38908 |
| Grey Matter mm3 (mean (SD)) | 616074 (55455) | 601073 (56175) | 38911 |
| WMH mm3 (median (IQR)) | 2708 (3936) | 4058 (6175) | 37168 |
| gFA units M (SD) | 0.01 (0.55) | -0.10 (0.58) | 34718 |
| gMD units M (SD) | -0.01 (0.46) | 0.12 (0.50) | 34718 |
| Ventricular CSF mm3 (mean (SD)) | 35817 (15847) | 41925 (17708) | 38721 |
| Hippocampus mm3 (mean (SD)) | 3843 (433) | 3755 (446) | 38876 |
| Accumbens mm3 (mean (SD)) | 443 (105) | 406 (103) | 38902 |
| Amygdala mm3 (mean (SD)) | 1245 (216) | 1247 (218) | 38896 |
| Pallidum mm3 (mean (SD)) | 1779 (221) | 1727 (233) | 38835 |
| Putamen mm3 (mean (SD)) | 4801 (569) | 4694 (571) | 38872 |
| Caudate mm3 (mean (SD)) | 3473 (419) | 3421 (419) | 38870 |
| Thalamus mm3 (mean (SD)) | 7667 (727) | 7465 (729) | 38851 |
| Pairs Matching - incorrect matches (mean (SD)) | 3.65 (2.86) | 3.82 (3.09) | 35898 |
| Verbal and Numerical Reasoning – Correct answers (mean (SD)) | 6.65 (2.05) | 6.27 (2.13) | 35837 |
| Reaction Time in seconds (median (IQR)) | 574 (129) | 593 (137) | 36323 |
| Trail-Making Test B – A in seconds (median (IQR)) | 292 (197) | 326 (235) | 24402 |
| Matrix Reasoning – Correct answers (mean (SD)) | 8.02 (2.12) | 7.53 (2.28) | 25286 |
| Symbol-Digit Substitution – Correct answers (mean (SD)) | 19.1 (5.23) | 17.2 (5.36) | 25320 |
| Tower Rearranging – Correct answers (mean (SD)) | 9.93 (3.22) | 9.54 (3.37) | 25074 |

Townsend Deprivation split into quintiles where 5 is most deprived; IQR:Interquartile range. For the brain measures larger values for white matter hyperintensities (WMH), ventricular CSF and g Mean Diffusivity (gMD) indicate poorer brain health whereas for all other brain measures smaller values
indicate poorer brain health. For the cognitive tests lower values indicate poorer cognition for verbal and numerical reasoning, matrix reasoning, symbol digit substitution and tower arranging and for pairs matching, reaction time and trail-making test B-A higher values indicate poorer cognition.

The demographics of participants stratified by diabetes and hypertension diagnosis can be found in the supporting information (**Supporting Information Table 3**). Those with both diabetes and hypertension (n = 1283) were more likely to be male, without a degree, higher BMI, an ex-smoker and with high cholesterol. Those with just diabetes (n = 760) were younger and more likely not to be from Caucasian ethnicities and were more likely to be current smokers. Those with hypertension only (n = 9649) tended to have slightly fewer or comparable comorbidities with those with diabetes or with both diseases but had a higher prevalence compared to those with no diabetes or hypertension.

### Association between diagnosed diabetes and neuroimaging and cognitive outcomes

Individuals with diabetes are associated with poorer brain measures compared to those without diabetes, with the exception of the amygdala subcortical region (**Figure 1 Model 1**). However, when adjusting for cardiovascular related variables associations were attenuated with gFA, a measure of white matter integrity, no longer different between those with and without diabetes (**Figure 1 Model 2**).

**Figure 1:**
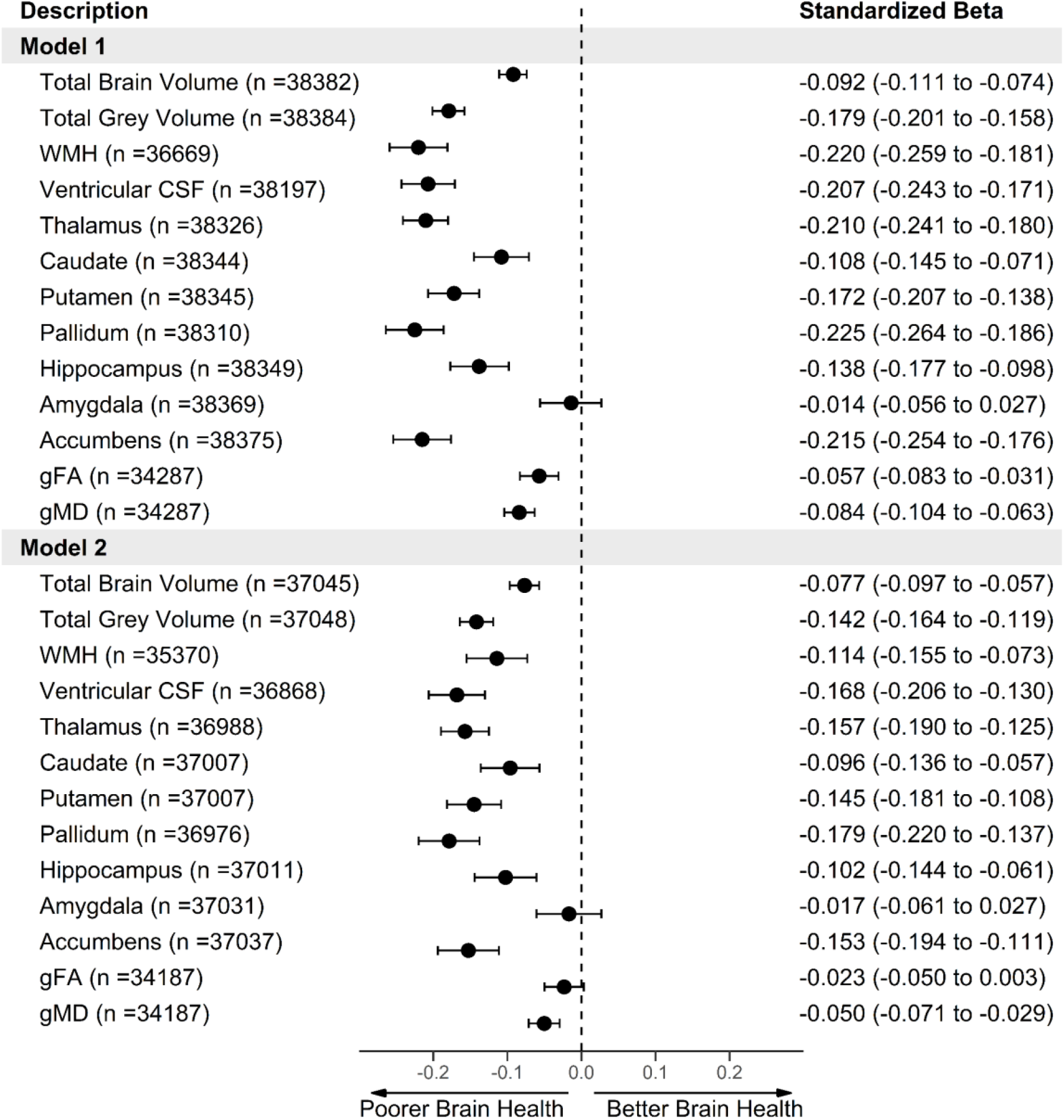
Forest plot of the association between diabetes (n=2043 cases, n=36875 controls) and neuroimaging outcomes. Model 1 = adjusted for age + sex + deprivation + ethnicity + educational attainment + head size + scanner position variables. Model 2 = Model 1 + BMI + CVD + hypercholesterolaemia + hypertension + smoking. WMH, Vascular CSF, gMD results were converted to the same direction of all other brain measures so that higher values indicate better brain health compared to reference level, for ease of comparisons.

When we compared Individuals with and without diabetes using the cognitive tests we found that in the partially adjusted models (model 1), those with diabetes had slower reaction times, slower on the trail making tests, poorer verbal and numeric reasoning and performed worse on matrix pattern and symbol digit substitution. There were no differences between groups for the pairs matching or tower rearranging (**Figure 2 Model 1**). Upon further adjustment for cardiovascular related covariates (model 2), results were again attenuated particularly for the matrix pattern cognitive and symbol digit substitution tests when comparing standardized betas between models 1 and 2 (**Figure 2 Model 2**).

**Figure 2:**
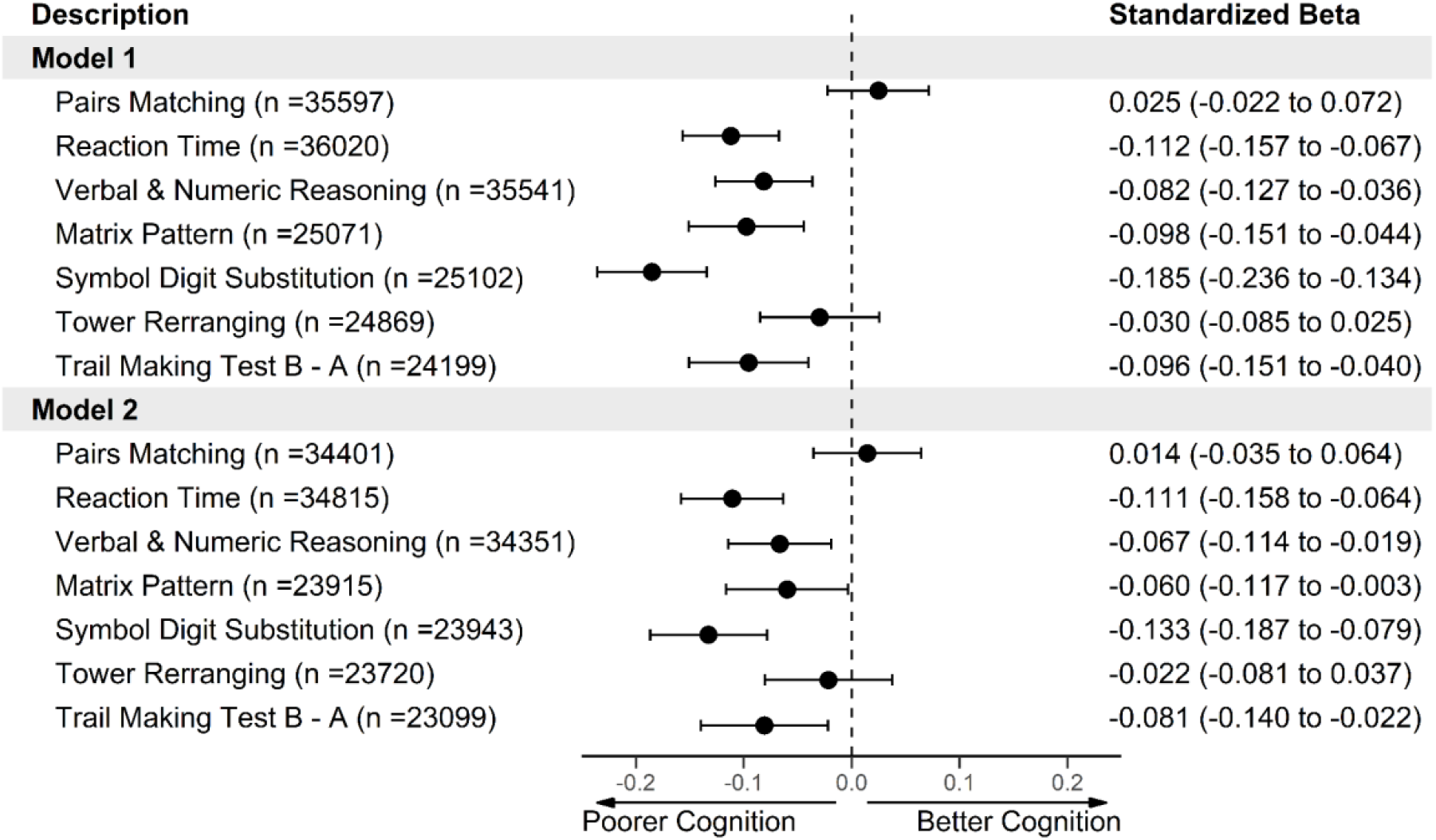
Forest plot of the association between diabetes (n=2043 cases, n=36875 controls) and cognitive function. Model 1 = adjusted for age + sex + deprivation + ethnicity + educational attainment. Model 2 = Model 1 + BMI + CVD + hypercholesterolaemia + hypertension + smoking. Reaction time, pairs matching and Trail making test B-A results were converted to the same direction of all other cognitive tests so that higher values indicate better cognitive performance and lower values indicate poorer cognitive performance compared to controls.

### Association between comorbid diabetes and hypertension vs only diabetes/hypertension vs neither: neuroimaging and cognitive outcomes

To investigate the interrelationship between comorbid diabetes and hypertension and neuroimaging and cognitive outcomes we further split our sample and created four groups, based on both diabetes and hypertension diagnosis. Using those individuals with both diabetes and hypertension as the reference group allowed the comparison of neuroimaging and cognitive outcomes with individuals with either diabetes or hypertension or neither disease to understand how having both diseases associated with brain and cognitive health.

**Figure 3** shows the fully adjusted (model 2) results of comparing individuals with comorbid diabetes and hypertension with those with either hypertension or diabetes and those not with these diseases with neuroimaging outcomes. The results for model 1, only adjusted for demographics, can be found in the supporting information **Figure S2**. From **Figure 3**, for the vast majority of neuroimaging outcomes, those with hypertension had better brain measures (i.e. larger brain volumes, smaller WMH) compared to those with both diabetes and hypertension apart from amygdala and gFA. Individuals with diabetes had larger total grey matter but there were no differences between individuals with diabetes only versus those with both hypertension and diabetes for total brain volume and subcortical volumes apart from the accumbens. Those with diabetes had lower WMH and better white matter integrity measures (gFA and gMD). Apart from the amygdala, those with neither disease had better brain health compared to those with both diseases.

**Figure 3:**
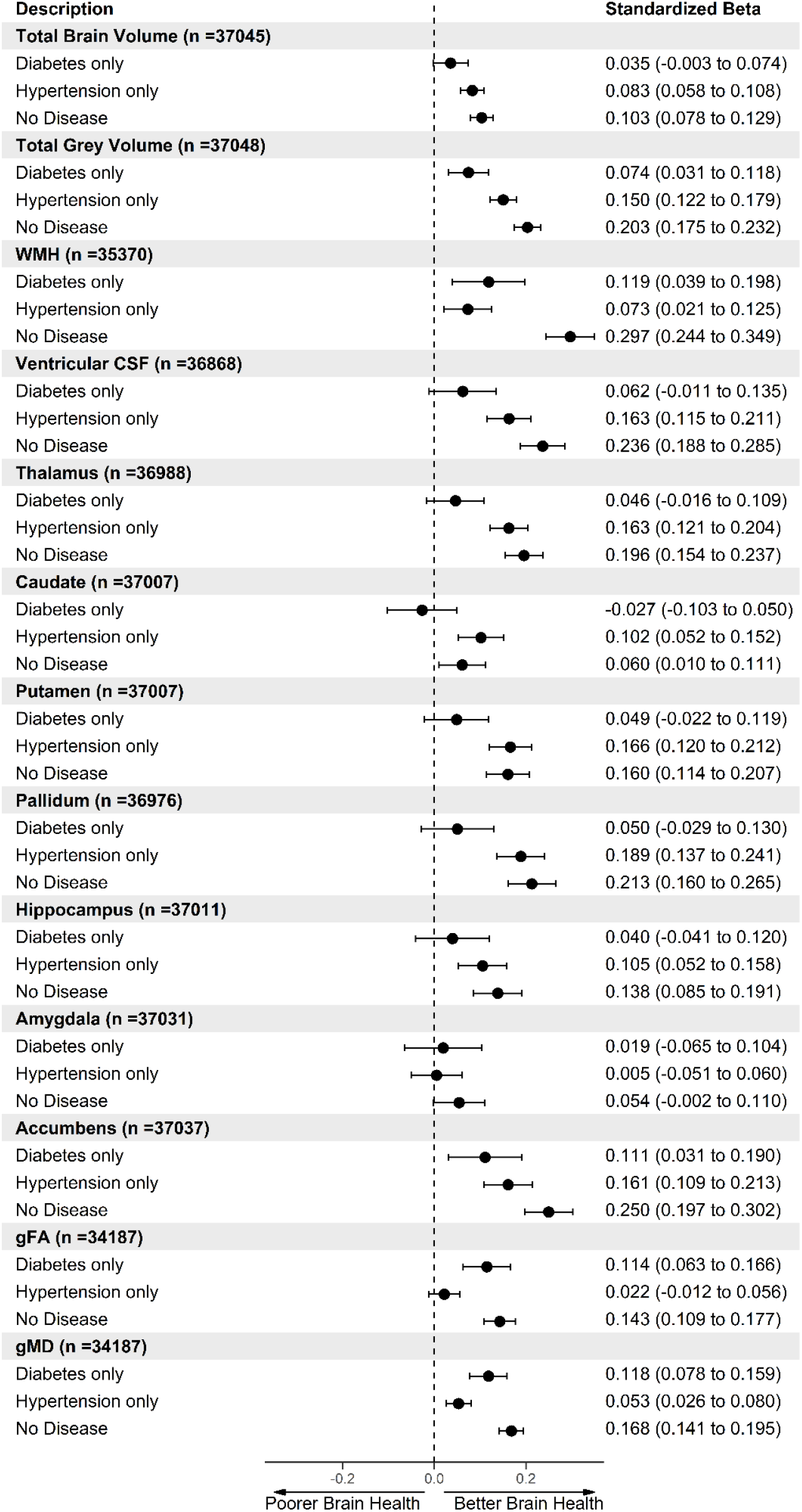
Forest plot of the association between disease status [both diabetes and hypertension (n = 1283), diabetes only (n = 760), hypertension only (n = 9649) and no diabetes and hypertension (n = 27226)] and neuroimaging outcomes. Individuals with both diabetes and hypertension were set as the reference level. Model 2 = adjusted for age + sex + deprivation + ethnicity + educational attainment + head size + scanner position variables + BMI + CVD + hypercholesterolaemia + smoking. WMH, Vascular CSF, gMD results were converted to the same direction of all other brain measures so that higher values indicate better brain health compared to reference level, for ease of comparisons.

Next, we carried out the same analysis as above instead using the cognitive test outcomes. The results for model 2 for the cognitive tests are presented in **Figure 4** (model 1 results can be found in Supporting information **Figure S3**). The results from **Figure 4** show those with hypertension or no disease had faster reaction times and performed better on the symbol digit substitution test. Individuals without hypertension and diabetes also performed better on the matrix pattern and Trail making tests. Interestingly those with diabetes only performed worse on the verbal and numerical reasoning test compared to individuals with both diabetes and hypertension.

**Figure 4:**
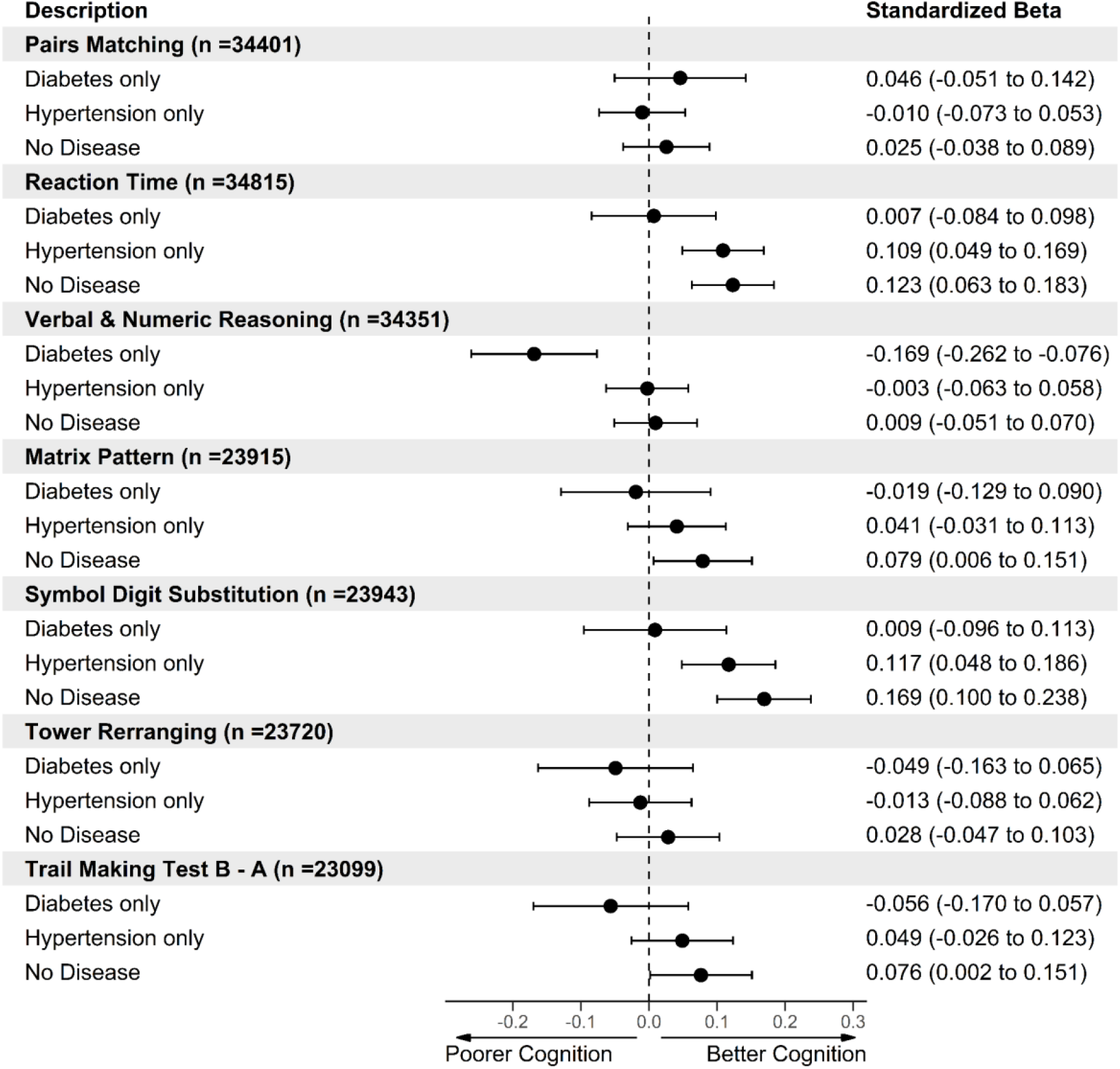
Forest plot of the association between disease status [both diabetes and hypertension (n = 1283), diabetes only (n = 760), hypertension only (n = 9649) and no diabetes and hypertension (n = 27226)] and cognitive function. Individuals with both diabetes and hypertension were set as the reference level. Model 2 = adjusted for age + sex + deprivation + ethnicity + educational attainment + BMI + CVD + hypercholesterolaemia + smoking. Reaction time, pairs matching and Trail making test B-A results were converted to the same direction of all other cognitive tests so that higher values indicate better cognitive performance and lower values indicate poorer cognitive performance compared to controls.

## DISCUSSION

Using data from a large population-based cohort, aged between 40 and 69 years, we aimed to investigate the complex interrelationships between diabetes, hypertension, neuroimaging measures and cognitive function. The main findings of our study are twofold. Firstly, individuals with diabetes have overall poorer brain and cognitive health, as indexed by global, subcortical and white matter regions and a range of cognitive tests. Secondly, individuals with both diagnosed diabetes and hypertension have poorer cognitive and brain health in comparison with those with hypertension only, and to a lesser extent those with only diabetes.

Individuals with diabetes had overall poorer brain health, as indexed by a range of volumetric brain parameters. These associations were present in our fully adjusted models for all neuroimaging outcomes, except for gFA (a measure of white matter integrity) and the amygdala (region associated with emotional processing and fear-related memories). These findings largely agree with previous studies of the association between diabetes and neuroimaging outcomes in the UKB [6, 7]. A key difference between our study and that of Cox et al. though, is that we observed no relationship between diabetes and gFA, but they did. This discrepancy could be due to differences in modelling approaches (including covariate adjustment), sample size (∼10,000 vs our sample of maximum ∼40,000) and definition of diabetes (self-reported vs use of a validated algorithm plus HbA1c measurements). Our study also investigated a much wider range of neuroimaging parameters compared to at least one of the earlier studies [7], which only included hippocampal and white matter hyperintensity volumes.

Similarly, we observed that those with a diabetes diagnosis also had poorer performance on certain cognitive function tests, particularly for reaction time, verbal and numeric reasoning, trail making and symbol digit substitution tests. Our finding for reaction time is in line with two previous UKB studies [5, 7] and the verbal numeric reasoning result is supported by one earlier study [5]. More broadly, it has long been suggested that individuals with diabetes have slower reaction times. Almost forty years ago, Subramanian and Chandrasekar showed that participants with diabetes had slower reaction times than age-matched controls [17].

Taken together, our findings of an association between diabetes and certain neuroimaging outcomes, as well as specific cognitive tests, warrant discussion. These associations may indicate that diabetes has a differential impact on brain regions and that for example, it has little impact on the amygdala and that perhaps emotional processing is not impaired in diabetes. However, experimental studies in mice do suggest that diabetes dysregulates dopaminergic circuitry in the amygdala and that this negatively affects certain aspects of social behavior [18]. In relation to diabetes and gFA, it is also possible that when taking into account other important factors (fully adjusted models) diabetes may not have a large impact on white matter integrity in mid-life. However, this should be interpreted with caution because as mentioned above, an earlier UKB study suggested that diabetes does relate to poorer white matter integrity [6]. A recent systematic review in fact suggests that findings of the relationship between diabetes and white matter integrity are equivocal, with some studies finding an association and others that do not [19].

Perhaps our most novel and intriguing finding is the strong association between comorbid diabetes and hypertension, and worse brain and cognitive health. Those with both conditions had worse overall brain health as measured by structural MRI and as expected, individuals without either diabetes/hypertension had better brain health. There was no difference between those with only diabetes vs those with both diabetes and hypertension for: total brain volume, ventricular CSF, thalamus, caudate, putamen, pallidum, hippocampus, amygdala. However, the largest difference was observed between those with only diabetes and those with both diseases in terms of total grey matter volume, a finding which agrees with a meta-analysis of this association [20]. Specifically, amongst the seven studies included in the meta-analysis, individuals with diabetes had reduced grey matter, compared with those without diabetes.

Having only hypertension vs both conditions was not associated with differences in the amygdala or gFA (white matter integrity). These results suggest that hypertension alone is with better brain health across a breadth of brain regions in comparison with only having diagnosed diabetes. The largest difference we observed between hypertension and comorbid diabetes/hypertension was in the pallidum, a structure within the basal ganglia which plays an important part in reward processing [21]. It is also important to note that when we examined white matter hyperintensity volume more closely (a marker of vascular brain damage) the size of the coefficient also suggested that hypertension related more strongly to this outcome in comparison with diabetes.

For cognitive function, the pattern of results was very similar to the brain imaging outcomes. Having only diabetes, in comparison with both diabetes and hypertension conferred worse performance on the verbal and numerical reasoning task, but no other cognitive tests. Importantly, comorbid diabetes and hypertension was associated with poorer performance on the reaction time and symbol digit substitution (processing speed/executive function) tasks, in comparison with having only hypertension, with very similar effect sizes across both of these tests.

Finding that comorbid diabetes and hypertension associates with worse overall brain and cognitive health may be novel, but it is not surprising. Epidemiological research that is almost two decades old suggests that those with both conditions have an increased dementia risk [22]. Our study indicates that comorbid diabetes and hypertension is strongly related to more objective, subclinical markers of dementia, as indexed by cognitive tests and structural MRI scans. From a clinical perspective, our findings suggest that perhaps there could be benefits for brain and cognitive health of using medication to treat both diabetes and hypertension in synergy. Substantial trial evidence shows that this has certainly been the case for microvascular and macrovascular complications [23]. However, one major difference is that there is uncertainty about whether the effects that we observed in our study are causal, in nature.

Strengths of our study include: the large sample of participants with data on cardiovascular risk factors, cognitive tests and particularly, structural brain MRI scans; the use of a validated algorithm and HbA1c levels to define prevalent diabetes [24]; and that we analysed standardised cognitive tests that were administered at the imaging visit in the UKB. However, we also acknowledge some important limitations: the UKB had a low response rate, which may result in selection bias [25]; the use of self-reported data for certain medical diagnoses, which could lead to misclassifications; and the use of observational data, which precludes causal conclusions.

In terms of the generalisability of our findings, we observed strong associations between diabetes and brain health even in relatively healthy participants in the UKB, which could mean that these effects would be more pronounced in a sample with greater external validity. The fact that our sample is largely white European also restricts the generalisability to other ethnic groups.

In conclusion, diabetes is associated with worse overall global brain health, as measured by structural MRI and not only those strongly related to dementia and cognitive decline (for example, the hippocampus). Our findings show that we should place greater emphasis on trying to understand when these detectable differences in brain structure might occur, as this could prove to be crucial in terms of dementia progression in individuals with diabetes. Moreover, we show that individuals with both hypertension and diabetes have worse overall brain and cognitive health when compared to those with only one of these diseases. This finding is novel and key to cardiovascular risk factor management for brain and cognitive health, more broadly. Overall, our research suggests that prevention of both diabetes and hypertension may delay structural brain changes, as well as cognitive decline and dementia.

## Data Availability

UK Biobank is an open access resource available to verified researchers upon application (http://www.ukbiobank.ac.uk/). Analysis syntax is available on request. This research was conducted using the UK Biobank Resource under approved project 43309.

## ABBREVIATIONS

AD: Alzheimer’s disease
BMI: body mass index
FA: Fractional anisotropy
MD: Mean Diffusivity
WMH: white matter hyperintensities

## Acknowledgments

We thank the UK Biobank participants and the UK Biobank team for their work in collecting, processing, and disseminating these data for analysis. We thank John Isaac (Janssen Pharmaceuticals), Professor Lenore Launer (National Institute of Aging), Associate professor Alejo Nevado-Holgado (University of Oxford) and Dr Laura Winchester (University of Oxford) for initial discussions. We also thank Associate Professor Rohini Mathur (LSHTM) for intellectual input.

## Author contributions

DN and VG both conceived and designed the study. DN carried out the analysis and both authors wrote the manuscript.

## Role of the funding source

The funding sources has no role in the study design; in the collection, analysis, and interpretation of data; in the writing of the report; and in the decision to submit the paper for publication.

Both authors accept full responsibility for the work and DN had access to the data, and both authors controlled the decision to publish.

## Supporting Information

**Table S1.** Self-reported health variables codes used for exclusion criteria on initial population.

| Condition | Code |
| --- | --- |
| *Self reported Illness (Field ID 20002)* |  |
| Dementia or Alzheimer's disease | 1263 |
| Parkinson's disease | 1262 |
| Chronic degenerative neurological | 1258 |
| Guillain-Barré syndrome | 1256 |
| Multiple Sclerosis | 1261 |
| Other demyelinating disease | 1397 |
| Brain cancer | 1032 |
| Brain haemorrhage | 1491 |
| Brain/intracranial abscess | 1245 |
| Cerebral aneurysm | 1425 |
| Cerebral palsy | 1433 |
| Encephalitis | 1246 |
| Epilepsy | 1264 |
| Head injury | 1266 |
| Infections of the nervous system | 1244 |
| Meningeal cancer | 1031 |
| Meningioma (benign) | 1659 |
| Meningitis | 1247 |
| Motor Neuron Disease | 1259 |
| Neurological injury/trauma | 1240 |
| Spina bifida | 1524 |
| Subdural haematoma | 1083 |
| Subarachnoid haemorrhage | 1086 |

**Table S2.** UK Biobank Field codes for all variables used in manuscript.

| Variable | Code |
| --- | --- |
| **Type 2 Diabetes variables** |  |
| Self reported diagnosis | Field ID 20002, code 1223 |
| Self reported anti diabetic drug medications | Field ID 20002, code 1140874646, 1140874664, 1140874674, 1140874686, 1140874706, 1140874718, 1140874744, 1140884600, 1141189090, 1141171646, 1140868902, 1141168660, 1141152590, 1140883066, 1140874650, 1140874652, 1140874658, 1140874660, 1140874666, 1140874678, 1140874680, 1140874690, 1140874712, 1140874716, 1140874724, 1140874726, 1140874728, 1140874732, 1140874736, 1140874740, 1140874746, 1141177600, 1141171652, 1140868908, 1141168668, 1141157284 |
| First occurrence | 130709 |
| HbA1c | 30750 |
| Diabetes diagnosed by a doctor | 2443 |
| Age of diabetes diagnosed | 2976 |
| **Hypertension variables** |  |
| Self reported diagnosis | Field ID 20002, Code 1065, 1072 |
| Age hypertension diagnosed | 2966 |
| BP medication use | Field IDs 6177, 6153 |
| **Neuroimaging** |  |
| Total Brain Volume (TBV) | 25010 |
| Total Grey Matter (TGM) | 25006 |
| Total White Matter (WM) | 25008 |
| White matter hyperintensities (WMH) | 25781 |
| Ventricular CSF | 25004 |
| Hippocampus (L+R) | 25019/20 |
| Thalamus (L+R) | 25011/12 |
| Caudate (L+R) | 25013/14 |
| Putamen (L+R) | 25015/16 |
| Pallidum (L+R) | 25017/18 |
| Amygdala (L+R) | 25021/22 |
| Accumbens (L+R) | 25023/24 |
| gFA (fractional anisotropy) | 25488-25514 |
| gMD (mean diffusivity) | 25515-25541 |
| **Cognitive Tests** |  |
| Symbol-Digit | 23324 |
| Matrix Reasoning | 6373 |
| Verbal and Numeric Reasoning | 20016 |
| Reaction Time | 20023 |
| Pairs Matching | 399 |
| TMT A | 6348 |
| TMT B | 6350 |
| Tower Rearranging | 21004 |
| **Confounding Variables** |  |
| Education | 6138 |
| Townsend deprivation index at recruitment | 189 |
| Smoking Status | 20116 |
| Gender | 31 |
| Age at Assessment | 21300 |
| Assessment Centre | 54 |
| BMI | 21001 |
| Ethnicity | 21000 |
| High Cholesterol | Field ID 20002, Code 1473 & Field IDs 6177, 6153 |
| Head size | 25000 |
| Scanner Position X | 25756 |
| Scanner Position Y | 25757 |
| Scanner Position Z | 25758 |
| Scanner Position | 25759 |

Field IDs obtained only for imaging visit apart from Ethnicity where baseline visit information was also used. For Hb1Ac values from baseline and second follow up were utilised.

### Definition of diabetes and hypertension phenotypes

Based on the field variables defined in Table S2 we created phenotypes for diabetes and hypertension based on various related variables. Further information on the specific variables is specified below.

#### Self-report

We used information from self reported disease status and self reported medication use for both diabetes and hypertension reported at the imaging visit from touchscreen questionnaires and nurse interviews.

#### Clinical

For diabetes diagnosis we used first occurrence variables provided by UK Biobank. This category contains data showing the ‘first occurrence’ of any code mapped to 3-character ICD-10.

The data-fields were generated by mapping:

1. Read code information in the Primary Care data (Category 3000),
2. ICD-9 and ICD-10 codes in the Hospital inpatient data (Category 2000),
3. ICD-10 codes in Death Register records (Field 40001, Field 40002), and
4. Self-reported medical condition codes (Field 20002) reported at the baseline or subsequent UK Biobank assessment centre visit

#### Biochemistry

HbA1c assays were performed using five Bio-Rad Variant II Turbo analysers, manufactured by Bio-Rad Laboratories, Inc. and employ a High Performance Liquid Chromatography (HPLC) method. A validation study ensured that the analysers underwent a multi-instrument comparison to ensure that they were in agreement4. More details are outlined in detail in the UK Biobank HbA1c protocol (Tierney A, Fry D, Almond R, et al. UK Biobank Biomarker Enhancement Project Companion Document to Accompany HbA1c Biomarker Data. 2018;1–8) .Available from: https://biobank.ndph.ox.ac.uk/showcase/showcase/docs/serum_hb1ac.pdf. For this work, any participant with an HbA1c greater than 48 mmol/mol was defined as undiagnosed diabetes and assigned to the diabetes group.

### Information on cognitive tests

Further information of the cognitive function tests can be found on the UKBiobank website (https://biobank.ndph.ox.ac.uk/showcase/label.cgi?id=100026).

#### Verbal and Numerical Reasoning

A task with thirteen logic/reasoning-type questions and a two-minute time limit was labelled as ‘fluid intelligence’ in the UK Biobank protocol but is now referred to as ‘verbal-numerical reasoning’; http://biobank.ctsu.ox.ac.uk/crystal/field.cgi?id=20016). The maximum score is 13.

#### Pairs matching

A visual memory test was administered, labelled ‘pairs-matching’ (http://biobank.ctsu.ox.ac.uk/crystal/label.cgi?id=100030). Participants were asked to memorize the positions of six card pairs, and then match them from memory while making as few errors as possible. Scores on the pairs-matching test are for the number of errors that each participant made; therefore, higher scores reflect poorer cognitive function. The Pairs matching task had two versions: 3-pair and 6-pair. We used 6-pair version for this work. For the pairs matching, values over 30 were capped at 30, and only participants who completed the task were included.

#### Reaction time

Participants completed a timed test of symbol matching, similar to the common card game ‘Snap’. (http://biobank.ctsu.ox.ac.uk/crystal/field.cgi?id=20023). The score on this task was the mean response time in milliseconds across trials, which contained matching pairs.

From 2016 at the imaging visit additional validated cognitive tests were administered including Matrix Pattern, Symbol-Digit Substitution, tower rearranging and Trail-Making Tests (TMT) B and A.

#### Trail Making Test A and B

In this work, we used TMT B – A. Subtracting TMT A from TMT B removes the individual variance in speed of response and is considered a useful tool in clinical practice for dementia. Individuals who scored >250 s for TMT B were excluded as well as participants with a TMT B - TMT A score less than 0 and greater than 150 seconds were also excluded.

#### Matrix Pattern

The participant was presented with a series of matrix pattern blocks with an element missing and asked to select the element that best completed the pattern from a range of displayed choices. https://biobank.ctsu.ox.ac.uk/crystal/label.cgi?id=501

#### Symbol-Digit Substitution

The participant was presented with one grid linking symbols to single-digit integers and a second grid containing only the symbols. They were then asked to indicate the numbers attached to each of the symbols in the second grid using the first one as a key. https://biobank.ctsu.ox.ac.uk/crystal/label.cgi?id=502. The number of symbol digit matches made correctly were used.

#### Tower rearranging

The participant was presented with an illustration of three pegs (towers) on which three differently-coloured hoops had been placed. They were then asked to indicate how many moves it would take to rearrange the hoops into another specific position. The number of correct puzzles was reported. https://biobank.ctsu.ox.ac.uk/crystal/label.cgi?id=503

**Figure S1:**
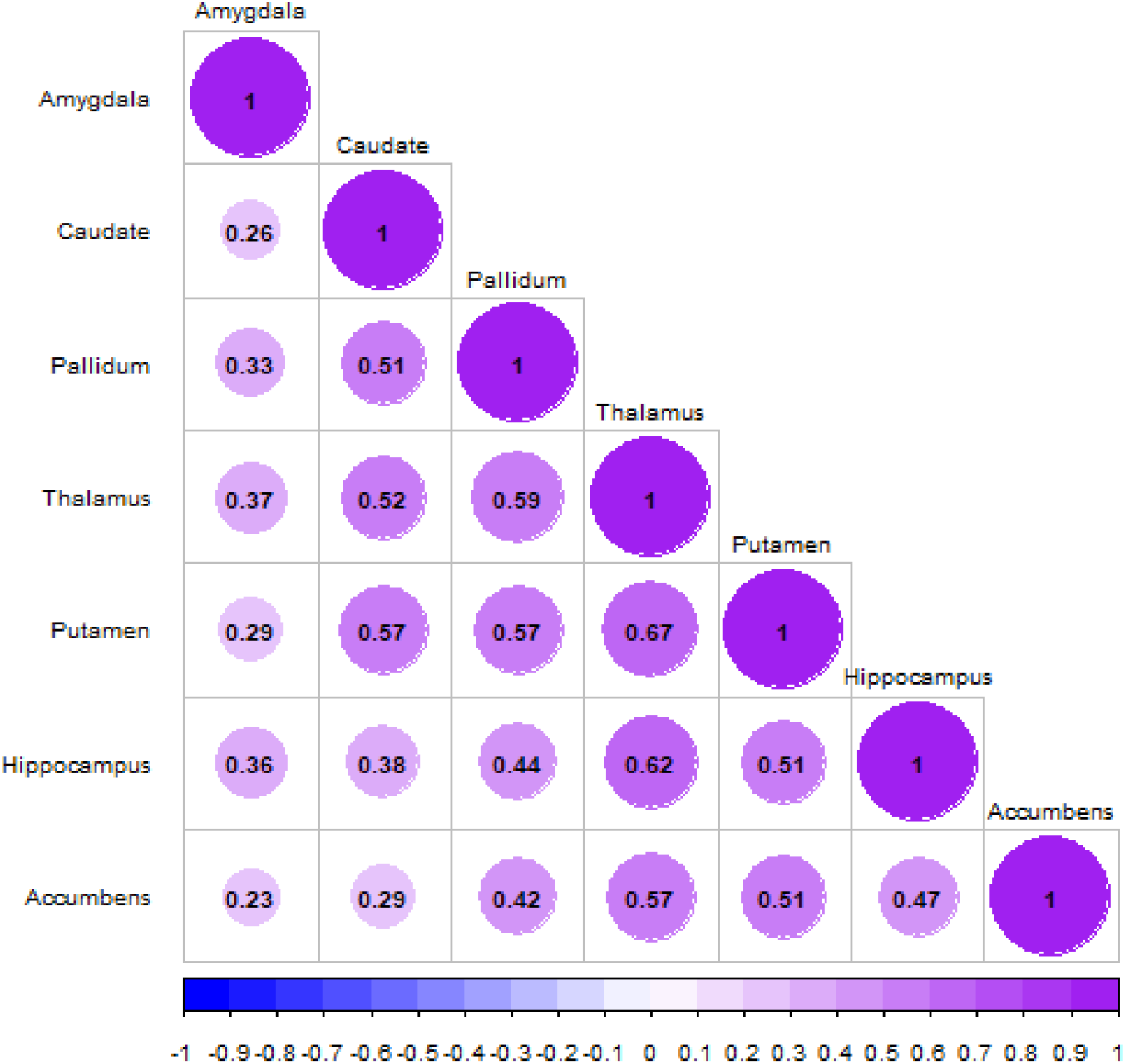
Pearson’s correlation matrix of subcortical regions brain measures. Higher values indicate higher correlation between regions.

**Table S3:** Characteristics of UK Biobank participants at imaging visit included stratified by diabetes and hypertension diagnosis.

| Description | No Disease (n=27226) | Hypertension (n = 9649) | Diabetes (n = 760) | Hypertension + diabetes (n = 1283) | N |
| --- | --- | --- | --- | --- | --- |
| Age years (mean (SD)) | 62.6 (7.52) | 66.1 (7.00) | 64.5 (7.49) | 66.6 (6.86) | 38918 |
| Gender (Male N (%)) | 11531 (42.4%) | 5499 (57.0%) | 464 (61.1%) | 844 (65.8%) | 38918 |
| Ethnicity (White N (%)) | 26422 (97.3%) | 9356 (97.2%) | 696 (91.8%) | 1179 (92.3%) | 38815 |
| Education (Degree N (%)) | 13797 (51.2%) | 4275 (44.7%) | 305 (40.6%) | 479 (37.8%) | 38525 |
| Townsend Deprivation (N (%)) |  |  |  |  |  |
| 1 | 5443 (20.0%) | 2011 (20.9%) | 116 (15.3%) | 219 (17.1%) | 38883 |
| 2 | 5549 (20.4%) | 1878 (19.5%) | 129 (17.0%) | 205 (16.0%) |  |
| 3 | 5403 (19.9%) | 1975 (20.5%) | 167 (22.0%) | 240 (18.7%) |  |
| 4 | 5456 (20.1%) | 1887 (19.6%) | 150 (19.7%) | 280 (21.9%) |  |
| 5 | 5348 (19.7%) | 1892 (19.6%) | 198 (26.1%) | 337 (26.3%) |  |
| Assessment Centre (N (%)) |  |  |  |  |  |
| Cheadle | 16926 (62.2%) | 6002 (62.2%) | 488 (64.2%) | 794 (61.9%) | 38918 |
| Reading | 3525 (12.9%) | 1245 (12.9%) | 90 (11.8%) | 162 (12.6%) |  |
| Newcastle | 6775 (24.9%) | 2402 (24.9%) | 182 (23.9%) | 327 (25.5%) |  |
| BMI Kg/m2 (mean (SD)) | 25.8 (4.04) | 27.8 (4.53) | 28.5 (4.97) | 30.5 (5.16) | 37626 |
| Smoking Status (N (%)) |  |  |  |  |  |
| Non Smoker | 17443 (64.7%) | 5655 (59.0%) | 417 (55.8%) | 675 (53.0%) | 38541 |
| Previous | 8527 (31.6%) | 3632 (37.9%) | 290 (38.8%) | 561 (44.1%) |  |
| Current | 973 (3.61%) | 291 (3.04%) | 40 (5.35%) | 37 (2.91%) |  |
| Hypercholesterolaemia (N (%)) | 3663 (13.5%) | 4402 (45.6%) | 416 (54.7%) | 1008 (78.6%) | 38918 |
| Hypertension (N (%)) | 0 (0.00%) | 9649 (100%) | 0 (0.00%) | 1283 (100%) | 38918 |
| Total Brain Volume mm3 (mean (SD)) | 1162722 (111172) | 1160787 (111180) | 1153345 (112068) | 1147126 (108711) | 38908 |
| Grey Matter mm3 (mean (SD)) | 617993 (55212) | 610659 (55783) | 606029 (56596) | 598138 (55738) | 38911 |
| WMH mm3 (median (IQR)) | 2394 (3276) | 3945 (5750) | 3448 (4644) | 4688 (7065) | 37168 |
| gFA units M (SD) | 0.05 (0.53) | -0.12 (0.59) | -0.01 (0.56) | -0.15 (0.58) | 34718 |
| gMD units M (SD) | -0.05 (0.43) | 0.13 (0.50) | 0.02 (0.46) | 0.17 (0.51) | 34718 |
| Ventricular CSF mm3 (mean (SD)) | 34368 (15119) | 39926 (17096) | 39642 (16811) | 43270 (18088) | 38721 |
| Hippocampus mm3 (mean (SD)) | 3857 (430) | 3804 (438) | 3779 (453) | 3740 (441) | 38876 |
| Accumbens mm3 (mean (SD)) | 450 (104) | 423 (104) | 420 (104) | 398 (102) | 38902 |
| Amygdala mm3 (mean (SD)) | 1244 (215) | 1249 (218) | 1244 (220) | 1249 (216) | 38896 |
| Pallidum mm3 (mean (SD)) | 1781 (219) | 1771 (228) | 1738 (213) | 1721 (244) | 38835 |
| Putamen mm3 (mean (SD)) | 4809 (564) | 4781 (583) | 4736 (561) | 4669 (575) | 38872 |
| Caudate mm3 (mean (SD)) | 3468 (416) | 3486 (426) | 3417 (417) | 3423 (421) | 38870 |
| Thalamus mm3 (mean (SD)) | 7690 (729) | 7602 (717) | 7516 (721) | 7434 (731) | 38851 |
| Pairs Matching - incorrect matches (mean (SD)) | 3.58 (2.81) | 3.87 (3.00) | 3.75 (3.28) | 3.86 (2.97) | 35898 |
| Verbal and Numerical Reasoning – Correct answers (mean (SD)) | 6.70 (2.05) | 6.53 (2.06) | 6.10 (2.06) | 6.37 (2.17) | 35837 |
| Reaction Time in seconds (median (IQR)) | 570 (125) | 585 (133) | 589 (120) | 597 (140) | 36323 |
| Trail-Making Test B – A in seconds (median (IQR)) | 285 (190) | 312 (216) | 323 (224) | 330 (245) | 24402 |
| Matrix Reasoning – Correct answers (mean (SD)) | 8.10 (2.10) | 7.76 (2.16) | 7.67 (2.31) | 7.45 (2.27) | 25286 |
| Symbol-Digit Substitution – Correct answers (mean (SD)) | 19.5 (5.17) | 18.0 (5.25) | 17.8 (5.26) | 16.9 (5.39) | 25320 |
| Tower Rearranging – Correct answers (mean (SD)) | 10.0 (3.22) | 9.64 (3.21) | 9.56 (3.32) | 9.53 (3.40) | 25074 |

Townsend Deprivation split into quintiles where 5 is most deprived, IQR:Interquartile range. For the brain measures larger values for white matter hyperintensities (WMH), ventricular CSF and g Mean Diffusivity (gMD) indicate poorer brain health whereas for all other brain measures smaller values indicate poorer brain health. For the cognitive tests lower values indicate poorer cognition for verbal and numerical reasoning, matrix reasoning, symbol digit substitution and tower arranging and for pairs matching, reaction time and trail-making test B-A higher values

**Figure S2:**
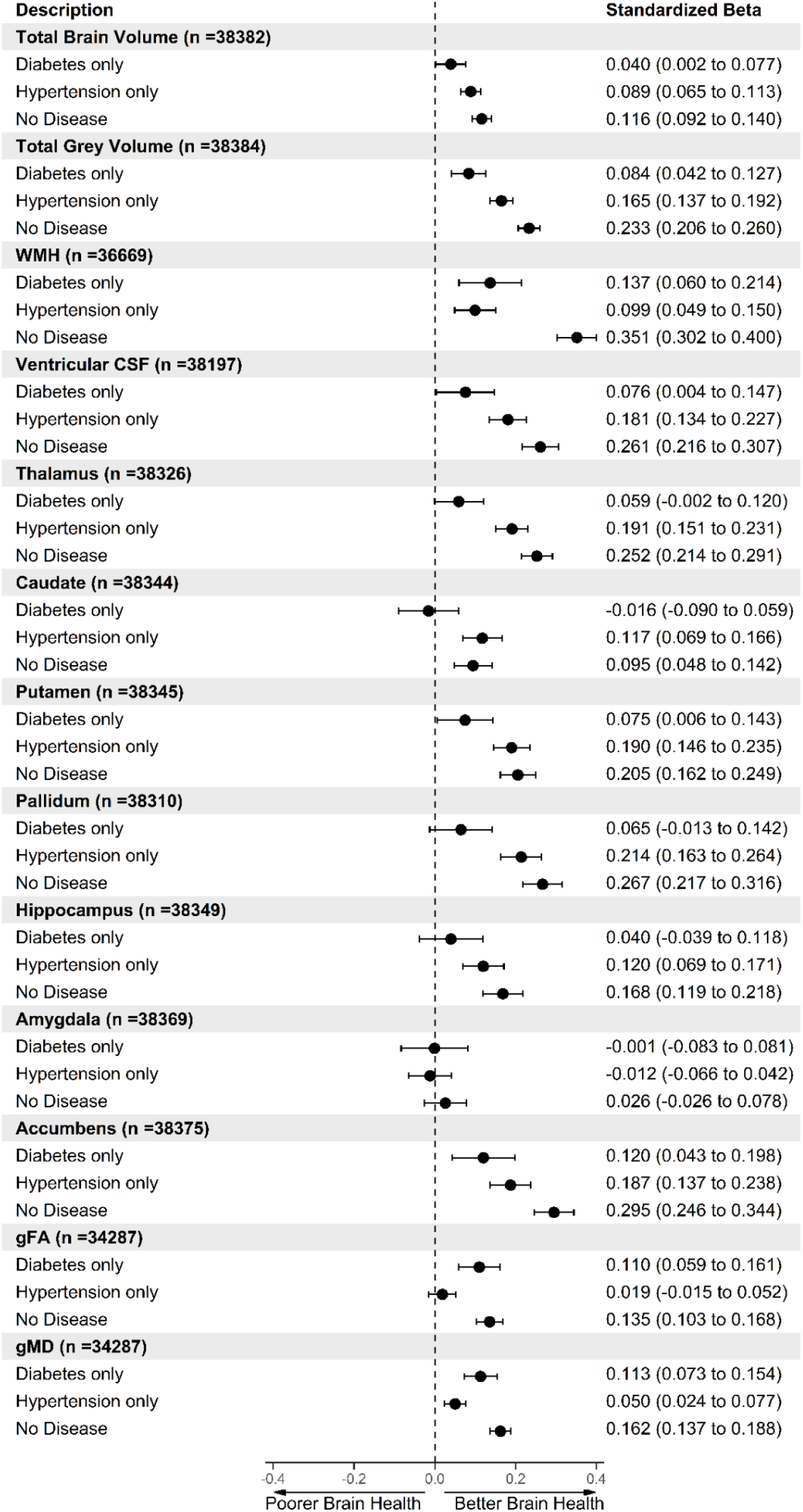
Forest plot of the association between disease status [both diabetes and hypertension (n = 1283), diabetes only (n = 760), hypertension only (n = 9649) and no diabetes and hypertension (n = 27226)] and neuroimaging outcomes. Individuals with both diabetes and hypertension were set as the reference level. Model 1 = adjusted for age + sex + deprivation + ethnicity + educational attainment + head size + scanner position variables. WMH, Vascular CSF, gMD results were converted to the same direction of all other brain measures so that higher values indicate better brain health compared to reference level for ease of comparisons.

**Figure S3:**
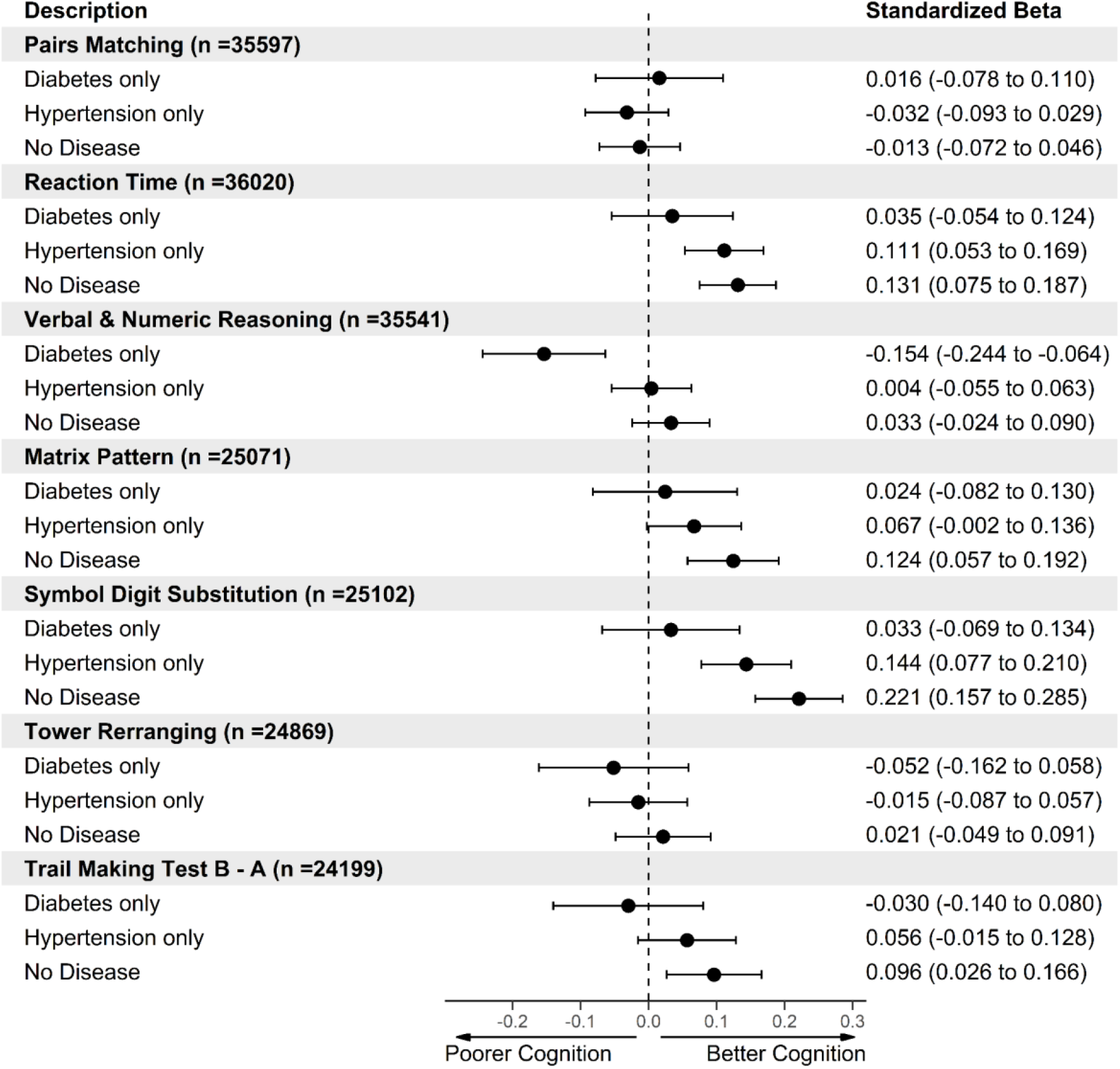
Forest plot showing the association of different cognitive measures between individuals with both diabetes and hypertension (n = 1283), diabetes only (n = 760), hypertension only (n = 9649) and no diabetes and hypertension (n = 27226). Individuals with both diabetes and hypertension were set as the reference level. Model 1 = adjusted for age + sex + deprivation + ethnicity + educational attainment. Reaction time, pairs matching and Trail making test B-A results were converted to the same direction of all other cognitive tests so that higher values indicate better cognitive performance and lower values indicate poorer cognitive performance compared to controls.

